# Stem Cell Therapies for Cartilage Defects: A Protocol for Systematic Review and Meta-Analysis of Randomized Controlled Trials

**DOI:** 10.1101/2024.05.23.24307771

**Authors:** Siddhartha Sharma, Ankit Dadra, Prasoon Kumar, Ashima, Riddhi Gohil, Sandeep Patel, Mandeep S. Dhillon

**Affiliations:** Foot and Ankle Biomechanics, Experimentation and Research (FABER) Laboratory, Department of Orthopedics, Postgraduate Institute of Medical Education and Research, Chandigarh, India; Department of Orthopedics, Fortis Hospital, Mohali, Punjab, India

**Author notes:** Correspondence to*: Dr. Siddhartha Sharma, Additional Professor, Department of Orthopedics, PGIMER, Chandigarh, India. 160012. Ethics Statement: Waiver of ethical clearance has been obtained for this systematic review and meta-analysis.

## Abstract

**Background:** Cartilage defects, arising from trauma, degenerative conditions, or genetic factors, pose significant challenges in orthopedic practice due to the limited regenerative capacity of articular cartilage. Traditional treatments like physical therapy and pain management often fail to restore tissue effectively. Recently, stem cell therapy has emerged as a promising regenerative medicine approach, potentially enhancing cartilage repair and improving joint function. This review investigates the efficacy of stem cell therapy in treating cartilage defects, focusing on pain reduction, disability, safety, quality of life, and cartilage regeneration compared to conventional care.

**Methods:** A PRISMA compliant systematic review and meta-analysis will be conducted by searching the PubMed, Embase, Web of Science, and Cochrane CENTRAL databases. Studies evaluating adult patients with cartilage defects of any etiology treated with various stem cell therapies will be included. Randomized controlled trials (RCTs) reporting these outcomes will be included, while non-randomized trials, non-comparative studies, non-human studies, and non-English publications will be excluded. Outcome measures will include validated functional outcome scores, rates of complications and adverse effects and healing of cartilage defects. Meta-analysis will be conducted for studies with similar interventions and outcomes, using fixed or random- effects models based on heterogeneity assessed by the I2 statistic. The quality of evidence will be assessed using GRADE criteria.

## 1. Introduction

Cartilage defects, resulting from trauma, degenerative conditions, or genetic predisposition, present a substantial challenge in modern orthopedic practice. The limited regenerative capacity of articular cartilage often leads to compromised joint function, chronic pain, and reduced quality of life for affected individuals. Traditional treatment approaches, such as physical therapy and pain management, aim to alleviate symptoms but often fall short in promoting true tissue restoration. In recent years, stem cell therapy has emerged as a promising avenue in regenerative medicine, offering the potential to address the underlying pathophysiology of cartilage defects. Stem cells, with their unique ability to differentiate into various cell types, hold the promise of restoring damaged cartilage and improving joint function [1]. This review explores the role of stem cell therapy in the treatment of patients with cartilage defects, examining its potential to enhance healing, reduce pain and disability, and ultimately revolutionize the management of a challenging orthopedic issue.

## 2. Research Question

What is the efficacy of stem cell therapy in treating patients with cartilage defects, as measured by reduction in pain, disability, safety (severe adverse events), quality of life, and cartilage regeneration, when compared to usual care/conventional care?

## 3. Inclusion and Exclusion Criteria

### 3.1 PICO Criteria

#### Patients

Adult patients with cartilage defects of any etiology (traumatic, osteoarthritis or idiopathic) in any location will be included.

#### Intervention

Any type of stem cell therapy (for e.g., umbilical cord derived stem cell therapy, bone marrow derived stem cell therapy, adipocyte derived stem cell therapy, peripheral blood derived stem cell therapy, stromal vascular fraction etc.) will be included. Standalone stem cell therapies, and therapies used in combination with procedures such as microfracture will be included.

#### Control

Any type of control therapy reported by the study authors will be included.

#### Outcomes

Validated functional outcome scores (such as VAS Score, NRS Score, IKDC Score, WOMAC Score etc.), rates and types of complications and healing of cartilage defects (ICRS or MOCART staging [2]) will be evaluated.

### 3.2 Inclusion Criteria

Randomized controlled trials, conducted on human subjects, evaluating the efficacy of any type of stem cell therapy or products derived from stem cells for treating cartilage defects. Studies should have reported validated functional outcome measures. There will be no restrictions on the year of publication.

### 3.3 Exclusion Criteria

- Nonrandomized controlled trials.
- Non-comparative studies (e.g., case reports, case series).
- Studies not reporting validated functional outcome measures.
- Non-English studies .
- Animal studies.

## 4. Search Strategy

Electronic searches will be conducted on PubMed, Embase, Web of Science and Cochrane CENTRAL databases.

Search terms will include variations of “stem cell therapy,” “cartilage defects,” and outcome measures (pain reduction, disability, safety, quality of life, functional assessment). Medical Subject Headings (MeSH) terms and relevant keywords will be used.

## 5. Study Selection

Two independent reviewers will screen titles and abstracts based on the inclusion/exclusion criteria. The Nested Knowledge platform will be used to manage references, screen studies and extract data for qualitative and quantitative synthesis . Full texts of potentially relevant studies will be retrieved and assessed by the same reviewers. Discrepancies will be resolved through adjudication of a third reviewer.

## 6. Data Extraction

Data extraction will be performed independently by two reviewers using a predefined data extraction form. Extracted data will include study characteristics (author, year, design), participant characteristics, intervention details (type of stem cell therapy), outcome measures, and results.

## 7. Risk of Bias Assessment

The risk of bias in included studies will be assessed using the Cochrane Risk of Bias 2 tool for RCTs [3]. Two reviewers will independently assess the risk of bias, and any disagreements will be resolved through discussion.

## 8. Data Synthesis and Analysis

Meta-analysis will be conducted for studies with similar interventions and outcome measures. Heterogeneity will be assessed using the *I*^*2 (*^I-squared) statistic. If *I*^*2*^ is found to be less than 50%, a fixed-effects model will be used to perform meta-analysis. If *I*^*2*^ is noted to be 50% or more, a random-effects model will be used. If numbers permit, subgroup analyses will be considered based on etiology of cartilage defect, route of administration of stem cell therapy and type of stem cell therapy used. The RevMan software will be used to perform meta-analyses.

## 9. Sensitivity Analysis and Publication Bias

Sensitivity analysis will be performed to assess the robustness of findings by excluding studies with high risk of bias. Publication bias will be assessed visually using funnel plots and quantitatively using methods such as Egger’s test.

## 10. Quality of Evidence

The quality of evidence will be assessed using GRADE (Grading of Recommendations Assessment, Development and Evaluation) criteria. The strength of evidence for each outcome will be categorized as high, moderate, low, or very low.

## 11. Reporting

The systematic review and meta-analysis will be reported following the Preferred Reporting Items for Systematic Reviews and Meta-Analyses (PRISMA) guidelines.

## 12. Ethical Considerations

Ethical approval waiver will be sought for this review as it will be based on published data.

## Data Availability

All data produced in the present study are available upon reasonable request to the authors

## List of Abbreviations

GRADE: Grading of Recommendations Assessment, Development and Evaluation
ICRS: International Cartilage Regeneration Society
MOCART: Magnetic Resonance Observation of Cartilage Repair Tissue
NRS: Numerical Response Scale
PRISMA: Preferred Reporting Items for Systematic Reviews and Meta-Analyses
RCT: Randomized Controlled Trial
VAS: Visual Analog Scale
WOMAC: Score Western Ontario and McMaster Universities Osteoarthritis Index

